# Detection of *Salmonella* Typhi and *bla*_CTX-M_ Genes in Drinking Water, Wastewater, and Environmental Biofilms in Sindh Province, Pakistan

**DOI:** 10.1101/2024.04.14.24305762

**Authors:** Ayesha Tajammul, L. Scott Benson, Jamil Ahmed, James VanDerslice, Windy Tanner

## Abstract

Typhoid fever poses a significant public health risk, particularly in low- and middle-income countries where access to clean water and improved sanitation may be limited. In Pakistan, this risk is especially serious given the emergence of an extensively drug-resistant (XDR) *Salmonella* Typhi strain, a strain attributed to *S*. Typhi acquisition of the *bla*_CTX-M-15_ gene. The now-dominant XDR *S*. Typhi strain, non-XDR *S*. Typhi, and *bla*_CTX-M_ genes are readily disseminated via drinking water and wastewater in Pakistan and may also be present in biofilms associated with these environmental sources. This study investigates the presence of *S*. Typhi and *bla*_CTX-M_ genes within these environmental compartments. Drinking water (n=35) or wastewater samples (n=35) and samples of their associated biofilms were collected from Karachi and Hyderabad, Pakistan. Samples were tested by PCR for *S*. Typhi and *bla*_CTX-M_ group 1 genes as a proxy for *bla*_CTX-M-15_. Heterotrophic plate counts (HPC) were conducted to assess microbial load. *S*. Typhi was detected by PCR in one bulk wastewater sample and one drinking water biofilm. *Bla*_CTX-M_ group 1 genes were detected in all sample types and were detected more frequently in bulk wastewater (n=13/35) than in drinking water (n=2/35) and more frequently overall in biofilm samples (n=22/70) versus bulk water (n=15/70). Detection of *bla*_CTX-M_ in biofilm was not significantly associated with detection in the associated bulk water sample. This study marks the first detection of *S*. Typhi in drinking water biofilms and the first report of *bla*_CTX-M_ genes in environmental biofilms in Pakistan. Environmental biofilms, particularly in drinking water systems, may serve as reservoirs for human exposure to *S*. Typhi and drug resistance genes. This study underscores the importance of expanding surveillance strategies to include biofilm sampling, providing valuable insights into pathogen dissemination in water systems, and informing targeted public health interventions to prevent waterborne diseases.

## Introduction

Typhoid fever is a serious cause of febrile illness that is more common in low- and middle-income countries (LMICs). The causative agent of typhoid fever is *Salmonella enterica* serotype Typhi (*S*. Typhi), for which humans are the only known animal reservoir [1]. Globally, there are an estimated 11 to 21 million cases of typhoid fever annually, resulting in 200,000 deaths [2]. Typhoid is primarily transmitted through consumption of food and water contaminated with *S*. Typhi bacteria that has been excreted by infected individuals or chronic carriers [2]; consequently, typhoid spread is higher in places lacking adequate sanitation or clean water access [3]. Pakistan is particularly vulnerable to typhoid fever outbreaks, as only about 20% of the population has access to clean drinking water, and an estimated 30% of all diseases and 40% of all deaths in Pakistan are attributed to poor water quality [4].

The advent of antibiotics has reduced the occurrence of typhoid fever [5]; however, indiscriminate antibiotic use has resulted in the emergence of multidrug-resistant (MDR) *S*. Typhi strains globally. More recently, an extensively drug-resistant (XDR) *S*. Typhi has emerged in Sindh, Pakistan. Between November 2016 and August 2021, 21,203 XDR typhoid fever cases were reported in Karachi, Hyderabad, and other districts of Sindh Province [6]. Currently, approximately 70% of typhoid fever cases in Sindh Province are classified as XDR [7], with the only remaining effective treatments being azithromycin (oral), carbapenems (parenteral), and tigecycline (parenteral) [8]. This XDR typhoid strain is believed to have arisen through the acquisition of a plasmid bearing a *bla*_CTX-M-15_ extended-spectrum beta-lactamase (ESBL) gene by *S*. Typhi, followed by clonal expansion [9]. Studies have reported that human carriage of *bla*_CTX-M-15_ is common in Pakistan [10] and prevalence in the environment is high [11-13].

Detection of *S*. Typhi in environmental samples using culture-based methods is notoriously difficult, and as a result, PCR detection has become the preferred method for detecting *S*. Typhi [14]. Environmental source type (e.g., soil, water, biofilm) could also affect *S*. Typhi detection. A recent study found that *S*. Typhi could be detected in naturally occurring biofilms on river rocks [14]. Biofilms are also an ideal venue for antimicrobial resistance gene transfer between microbes [15], which could be a critical concern if drinking water biofilms harbor serious pathogens and have a high prevalence of clinically important mobile resistance genes.

The primary objective of this study was to determine if *S*. Typhi could be detected in drinking water, wastewater, or their associated biofilms in urban areas in Sindh Province. Our second objective was to independently determine the environmental prevalence of the *bla*_CTX-M-15_ gene in drinking water, wastewater, and their associated biofilms.

## Materials and Methods

### Study Setting, sample collection, and filtration

The study was conducted in Sindh Province, the second most populated province of Pakistan’s four provinces, located in the southern-most part of the country. Sindh Province is a subtropical region and has an estimated population of 47.8 million people [4, 11]. The province includes two major cities, Hyderabad and Karachi, which have a primarily dry climate with hot summers and mild winters. Samples were collected in November 2020 and July 2021.

A total of 70 wastewater (n=35) or drinking water grab samples (n=35) were collected. Drinking water samples were collected from ground cisterns fed from the municipal piped water supply (n=30) or from outdoor taps drawing from ground water supplies at selected sites (n=5) in the greater Karachi and Hyderabad areas in Sindh Province, Pakistan (Figure 1a, 1b). Drinking water samples (1 liter) were collected in sterile Whirl-Pak bags (Whirl-Pak Filtration Group, Pleasant Prairie, Wisconsin, USA), and wastewater samples (50 - 100 mL total) were collected in sterile 50 mL conical tubes. In addition, biofilm samples associated with the water and wastewater sample sites (n=70) were collected by swabbing a 4 x 4 inch area of the water/wastewater pipe or ground cistern wall surface using a sterile ESwab regular swab with flocked nylon fiber tip (Copan Diagnostics, Murrieta, California, USA). Liquid wastewater and drinking water samples are referred to as bulk wastewater and bulk drinking water samples, whereas biofilm swab samples will be referred to as wastewater biofilm and drinking water biofilm samples.

**Figure 1.**
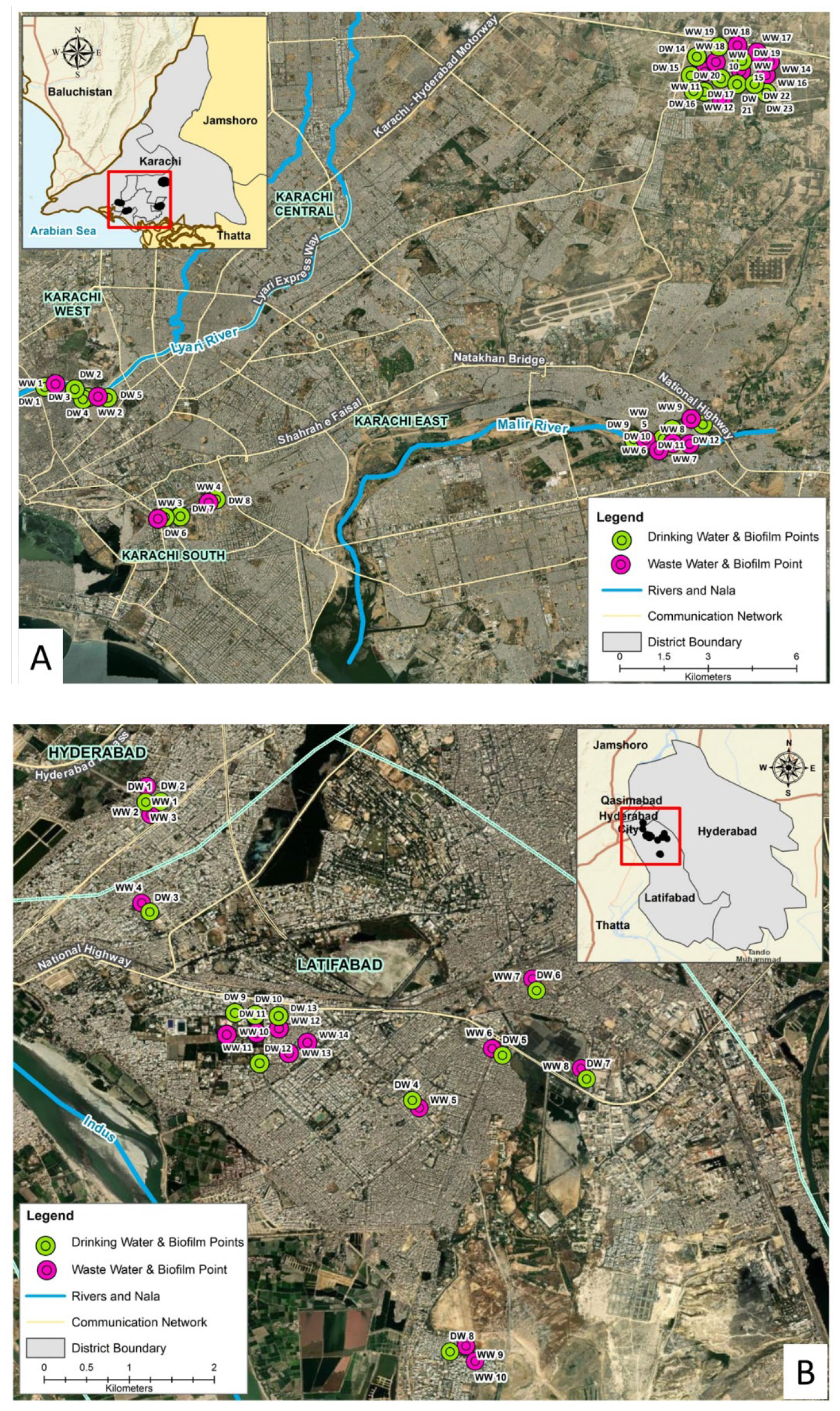
Map of sample collection points within Karachi (A) and Hyderabad (B) Pakistan for drinking water (DW) and wastewater (WW) and their corresponding biofilms.

Samples were transported in coolers with ice packs to the U.S.-Pakistan Center for Advanced Studies–Water (USPCAS-W), Mehran University of Engineering and Technology (MUET), Jamshoro, for testing. Wastewater samples were clarified by pre-filtering the samples through a 5 µm syringe filter to remove debris and larger organisms. Drinking water samples and the clarified wastewater samples were then filtered through 0.45 µM sterile disposable filter cups. Additional filters were used if the initial filter was clogged prior to passing through the entire measured volume. Filters were placed in 10 mL of sterile phosphate buffered saline and pulse vortexed. ESwabs were pulse-vortexed in the swab transport media, and the resuspended filtrate or biofilm was aliquoted onto heterotrophic plate count agar (Becton Dickenson, Franklin Lakes, New Jersey, USA) to determine a crude viable bacterial count per sample using a spot-titer culture assay method by plating 10 µL of 10-fold dilutions (10^-1^ to 10^-5^) [16].

### DNA extraction and PCR

DNA was extracted from an aliquot of resuspended PBS filtrate or ESwab transport medium using a PowerSoil DNA isolation kit (Qiagen, Germantown, Maryland, USA). Sample DNA extractions were tested at USPCAS-W, MUET by conventional PCR for the presence of *bla*_CTX-M-15_ genes and by quantitative PCR (qPCR) for *S*. Typhi genes. Subsequently, DNA-extracted and RNAse-treated samples were shipped to the Yale School of Public Health in the U.S. for confirmatory PCR testing and confirmation.

CTX-M group-1 primers were used as a proxy for the detection of the *bla*_CTX-M-15_ variant, as *bla*_CTX-M-15_ is the predominant CTX-M variant within CTX-M group 1 [17, 18]. Primer and probe sequences for CTX-M group-1 and *S*. Typhi were acquired from the Yale Keck Oligonucleotide Synthesis facility (New Haven, Connecticut, USA). A primer set for detection of CTX-M group 1 alleles was used as proxy for *bla*_CTX-M-15_ gene detection, as *bla*_CTX-M-15_. Extracted DNA from a clinical *S*. Typhi strain was used for the *S*. Typhi PCR conducted in Pakistan. For confirmatory PCR work conducted in the U.S., gBlocks gene fragments (Integrated DNA Technologies (IDT) Coralville, Iowa, USA) were used as positive amplification controls for *S*. Typhi and CTX-M group 1 based on representative reference sequences in GenBank (CP053702.1 for *S*. Typhi and KU355874.1 for CTX-M group 1) that fell between the forward and reverse primer targets. Primer and probe sequences can be found in Table 1 for the specific PCR assays. PCR reactions for *S*. Typhi and CTX-M group 1 were performed in single plex conventional PCR format using Hot Start Taq 2x Master Mix (New England Biolabs, Ipswich, Massachusetts, USA). PCR products were run on a 1% agarose gel.

**Table 1.**
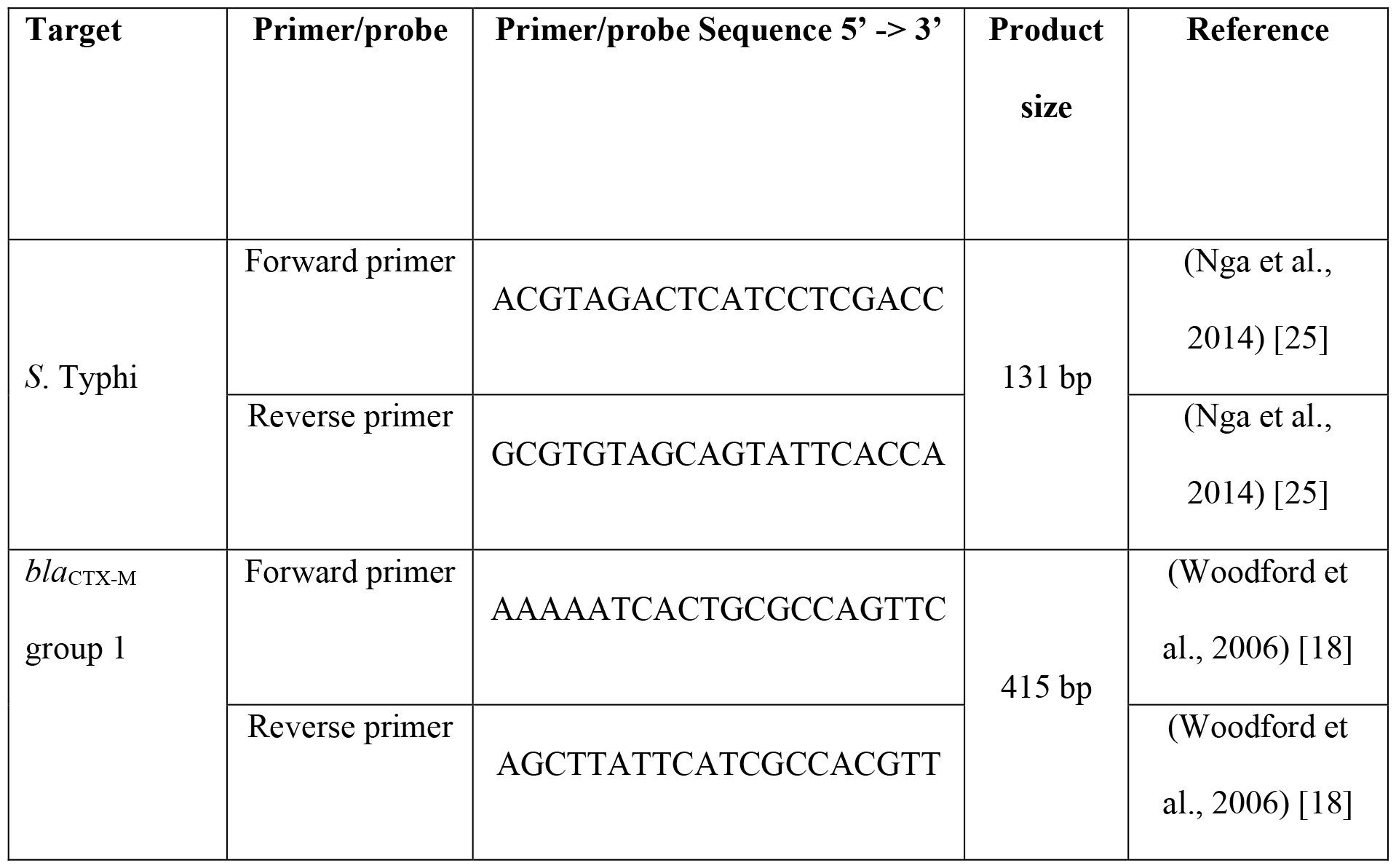
Primers used in conventional PCR to detect *S*. Typhi and *bla*_CTX-M_ group 1 genes.

## Results

*S*. Typhi DNA was detected in two of the 140 samples: in one bulk wastewater sample and one drinking water biofilm sample. The *bla*_CTX-M_ group 1 genes (proxy for *bla*_CTX-M-15_) were identified in two bulk drinking water samples and one drinking water biofilm sample. The *bla*_CTX-M_ group 1-positive drinking water biofilm sample was not from the same site as either of the two sites where CTX-M group 1 was found in the bulk drinking water. In wastewater, *bla*_CTX-M_ group 1 genes were identified in 13 bulk water samples as well as 21 wastewater biofilm samples. Of the 13 bulk wastewater samples positive for *bla*_CTX-M_ group 1 genes, 9 (69.2%) of the associated biofilm samples were also positive for *bla*_CTX-M_ group 1. Overall, 54.3% of the sample pairs were concordant, indicating no statistically significant association between the presence of *bla*_CTX-M_ group 1 genes in wastewater and the biofilm at the same sampling site (Table 2). The *bla*_CTX-M_ group 1 genes were not detected in either of the samples positive by PCR for *S*. Typhi (Figure 2).

**Table 2.** Number of water and biofilm samples in which the CTX-M group 1 gene was detected.

\* Indicates sample types in which one sample tested positive for *S. Typhi*.
|  | Drinking water | Wastewater | Total |
| --- | --- | --- | --- |
| Water | 2 (n=35) | 13 (n=35)* | 15 (n=70) |
| Biofilm | 1 (n=35)* | 21 (n=35) | 22 (n=70) |
| Total | 3 (n=70) | 34 (n=70) | 37 (n=140) |

**Figure 2.**
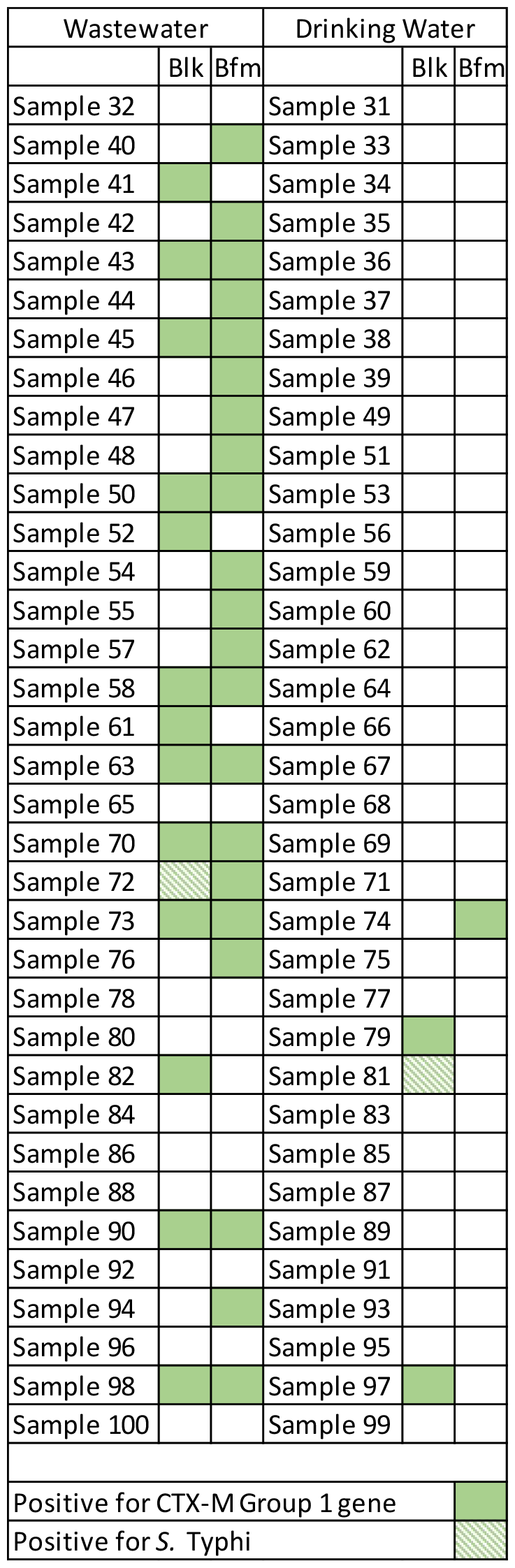
Bulk water and associated biofilm samples positive for *bla*_CTX-M_ group 1 genes (solid green) and *Salmonella* Typhi. Blk = bulk water sample; Bfm = biofilm sample.

Total heterotrophic plate counts (HPC) were higher in bulk waste water (n=35) compared to bulk drinking water (n=35) (median 2.9 x 10^8^ colony forming units [cfu] [IQR: 7.4 x 10^7^ cfu, 1.5 x 10^9^ cfu] vs. 1.5 x 10^5^ cfu [IQR: 7.3 x 10^4^ cfu, 6.7 x 10^5^ cfu]) and higher in wastewater biofilm (n=35) compared to drinking water biofilm (n=35) (2.0 x 10^7^ cfu [IQR: 1.0 x 10^7^ cfu, 5.0 x 10^7^ cfu] vs. median 2.0 x 10^5^ cfu [IQR: 3.0 x 10^4^ cfu, 7.5 x 10^6^ cfu]); HPC counts were similar between biofilm (n=70) and bulk water (n=70) samples (median 1.0 x 10^7^ colony forming units (CFU) [interquartile range (IQR): 1.3 x 10^5^ cfu, 3.0 x 10^7^ cfu] vs. 9.2 x 10^6^ cfu [IQR: 1.4 x 10^5^ cfu, 2.8 x 10^8^ cfu]) however, heterotrophic plate count comparisons between bulk water and biofilm samples were not informative, as bulk water was measured by volume and biofilm was measured by surface area, resulting in an arbitrary differences between the two sample types. Heterotrophic plate counts did not appear to differ significantly between samples in which *bla*_CTX-M_ was detected compared to samples in which it was not detected.

## Discussion

Environmental dissemination of increasingly drug-resistant *S*. Typhi to community water systems is a critical public health emergency. To our knowledge, this is the first study that has investigated the presence of *S*. Typhi in drinking water and wastewater biofilms and has identified *S*. Typhi in drinking water biofilms. It is also the first to demonstrate the widespread dissemination of *bla*_CTX-M_ group 1 genes in environmental biofilm in Pakistan. Our detections of *S*. Typhi in the bulk wastewater sample and the drinking water biofilm sample both occurred during the second sampling period in July 2021, with no detections during the November 2020 sampling period. The weekly National Institute of Health Islamabad Field Epidemiology Reports reported significantly more human cases of *S*. Typhi in Karachi in July 2021 than in November 2021 (80 cases vs. 14 cases) and an equal number of cases in Hyderabad (13 cases) [19, 20]; however, our detection of *S*. Typhi was too infrequent to make statistically significant associations with sampling period or environmental source (i.e. drinking water, wastewater, biofilm).

The detection of *S*. Typhi and CTX-M genes in biofilms, particularly in drinking water biofilms, has serious public health and environmental surveillance implications. Environmental surveillance for *S*. Typhi and other fecal-oral pathogens is important for determining microbial epidemiology and providing data for risk assessments and prevention of pathogen spread; however, current environmental surveillance strategies traditionally restrict sampling to bulk water, which may miss biofilms as an important environmental reservoir of *S*. Typhi and *bla*_CTX-M-15_. *S*. Typhi has previously been found in environmental biofilms [14], and *S*. Typhi recovered from the environment has demonstrated biofilm-forming capability [21]. This is highly relevant to *S*. Typhi dissemination via drinking water, as distribution systems are common substrates for environmental biofilm development. *S*. Typhi could also attach or become enmeshed in a distribution system biofilm and possibly even experience limited growth [22]. Changes to hydraulic flow could result in intermittent release of the biofilm and any associated *S*. Typhi into the water column, making it difficult to capture through routine collection of environmental surveillance samples [22].

Hyderabad and Karachi, as important metropolitan hubs in Sindh Province, confront considerable issues in providing adequate drinking water and sanitation amenities to their rising populations. Rapid urbanization, combined with insufficient investment in water and sanitation facilities, has left a significant percentage of the population reliant on contaminated water sources [4, 7]. Warmer temperatures can increase the prevalence of waterborne infections such as typhoid. Furthermore, the region’s erratic and intense rainfall makes it difficult to maintain clean water supplies and proper sanitation infrastructure. The combination of high temperatures, limited precipitation, and insufficient sanitation facilities promote *S*. Typhi persistence and transmission, particularly in wastewater and drinking water sources [4]. The contamination of drinking water with feces, which is frequently caused by the proximity of sewage lines to water supply systems or the introduction of pathogens into aged water pipelines, poses a serious danger of typhoid transmission. Furthermore, the creation of biofilms within water distribution networks hinders efforts to reduce bacterial contamination by providing a habitat for *Salmonella* Typhi to persist and multiply [6, 8].

In addition to *S*. Typhi detection, we determined the distribution and prevalence of *bla*_CTX-M-15_ genes in water, wastewater, and their associated biofilms, due to the importance of this gene in *S*. Typhi resistance to ceftriaxone in Pakistan. ESBL-producing Enterobacteriaceae have been found in water environments in many Asian countries and pose a substantial concern to public health due to their increased resistance to third generation cephalosporins (e.g., ceftriaxone, cefotaxime, and ceftazidime) as well as monobactams (aztreonam) [23]. We detected *bla*_CTX-M_ group 1 genes (as a proxy for the *bla*_CTX-M-15_ gene) in more than a quarter of the environmental samples. The *bla*_CTX-M_ group 1 genes were found significantly more frequently in bulk wastewater and biofilm (n=34) compared to bulk drinking water and biofilm (n=3). We also detected *bla*_CTX-M_ group 1 genes about 50% more frequently in biofilm samples (n=22) compared to bulk water samples (n=15), although these sample types are not directly comparable.

Bacteria harboring ESBL genes in drinking water, wastewater, and associated biofilms indicate a potential source of acquired resistance genes in humans. Although we did not determine the bacterial species associated with the *bla*_CTX-M_ genes, our study supports previous reports on ESBL-producing enterobacteria. A previous study found that ESBL-producing *E. coli* accounted for 57.7% (15/26) of *E. coli* isolates recovered in surface and wastewater in Islamabad, Pakistan [24]. Two other conference reports on the disseminations of *bla*_CTX-M_ genes among enterobacteria in water or wastewater in Faisalabad or Islamabad, Pakistan reported a prevalence of CTX-M-producing *E. coli* in 29% and 8% of samples, respectively [11, 12].

This study was limited by the number of time periods in which sampling was conducted and few *S*. Typhi-positive samples from which to draw conclusions about *S*. Typhi seasonality in environmental water and biofilms or parallel trends with human cases in Sindh, Pakistan. Other limitations included PCR primers that targeted the broader *bla*_CTX-M_ group 1 genes rather than *bla*_CTX-M-15_ specifically; however, a recent study of *E. coli* isolates from south Asia found that *bla*_CTX-M-15_ was the only *bla*_CTX-M_ variant detected among the *bla*_CTX-M_ group 1 isolates [17]. *S*. Typhi viability could not be ascertained utilizing a PCR assay for detection, and *bla*_CTX-M_ gene host could not be discerned through PCR testing of DNA from the whole environmental sample. Direct comparisons could not be made between *S*. Typhi or *bla*_CTX-M_ genes in biofilm versus bulk water as this would differ by arbitrary selection of bulk water volume versus biofilm surface area; therefore, we cannot definitively conclude which sample type yields better pathogen or resistance gene detection. Biofilm sampling also provides less certainty about the period when the pathogen or resistance gene passed through the system. The lack of concordance between detection of *S*. Typhi and *bla*_CTX-M_ in biofilms and the associated bulk water sample underscores the need for further investigation to better understand the information provided by biofilm sampling and its utility in identifying previous or current pathogen and resistance gene dissemination in drinking water and wastewater systems.

## Data Availability

The data supporting the findings of this study are available upon reasonable request from the corresponding author.

## Acknowledgements

This work was supported by the Bill & Melinda Gates Foundation Grant Number OPP1217099.

## References

1. Levine MM, Tapia MD, Zaidi AKM. CHAPTER 16 - Typhoid and Paratyphoid (Enteric) Fever. In: Guerrant RL, Walker DH, Weller PF, editors. Tropical Infectious Diseases: Principles, Pathogens and Practice (Third Edition). Edinburgh: W.B. Saunders; 2011. p. 121–7.

2. U.S. Centers for Disease Control and Prevention. Information for Healthcare Professionals 2023 [cited 2024 February 11.]. Available from: https://www.cdc.gov/typhoid-fever/health-professional.html.

3. World Health Organization. Typhoid 2024 [cited 2024 February 11]. Available from: https://www.who.int/news-room/fact-sheets/detail/typhoid.

4. Daud MK, Nafees M, Ali S, Rizwan M, Bajwa RA, Shakoor MB, et al. Drinking Water Quality Status and Contamination in Pakistan. BioMed Res Int. 2017;2017:7908183. doi: 10.1155/2017/7908183.

5. Das JK, Hasan R, Zafar A, Ahmed I, Ikram A, Nizamuddin S, et al. Trends, Associations, and Antimicrobial Resistance of Salmonella Typhi and Paratyphi in Pakistan. Am J Trop Med Hyg. 2018;99(3_Suppl):48–54. Epub 2018/07/27. doi: 10.4269/ajtmh.18-0145. PubMed PMID: 30047366; PubMed Central PMCID: PMCPMC6128361.

6. National Institute of Health (NIH) Islamabad. Weekly Field Epidemiological Report August 15-21, 2021. Field Epidemiology & Disease Surveillance Division; 2021 August 25, 2021. Report No.

7. Tanveer M, Ahmed A, Siddiqui A, Rehman IU, Hashmi FK. War against COVID-19: looming threat of XDR typhoid battle in Pakistan. Public health. 2021;198:e15–e6. Epub 2021/07/01. doi: 10.1016/j.puhe.2021.05.019. PubMed PMID: 34187703; PubMed Central PMCID: PMCPMC8139234.

8. Akram J, Khan AS, Khan HA, Gilani SA, Akram SJ, Ahmad FJ, et al. Extensively Drug-Resistant (XDR) Typhoid: Evolution, Prevention, and Its Management. Biomed Res Int. 2020;2020:6432580. Epub 2020/05/29. doi: 10.1155/2020/6432580. PubMed PMID: 32462008; PubMed Central PMCID: PMCPMC7212280.

9. Klemm EJ, Shakoor S, Page AJ, Qamar FN, Judge K, Saeed DK, et al. Emergence of an Extensively Drug-Resistant Salmonella enterica Serovar Typhi Clone Harboring a Promiscuous Plasmid Encoding Resistance to Fluoroquinolones and Third-Generation Cephalosporins. mBio. 2018;9(1). Epub 2018/02/22. doi: 10.1128/mBio.00105-18. PubMed PMID: 29463654; PubMed Central PMCID: PMCPMC5821095.

10. Habeeb MA, Haque A, Iversen A, Giske CG. Occurrence of virulence genes, 16S rRNA methylases, and plasmid-mediated quinolone resistance genes in CTX-M-producing Escherichia coli from Pakistan. European journal of clinical microbiology & infectious diseases: official publication of the European Society of Clinical Microbiology. 2014;33(3):399–409. Epub 2013/09/17. doi: 10.1007/s10096-013-1970-1. PubMed PMID: 24036893.

11. Ahad A, Salman M, Ikram A, Ashraf Z, Amir A, Saeed A, et al. Prevalence and molecular Characterization of ESBL-producing Escherichia coli in waste water samples from Pakistan. International Journal of Infectious Diseases. 2020;101:33. doi: 10.1016/j.ijid.2020.09.119.

12. Qamar MU, Rizwan M, Bashir I, Ali Q, Gilani MM, Saqalein M, et al. Prevalence and coexistence of mcr-1 and ESBL producing Escherichia coli isolated from poultry and environmental water from Pakistan. International Journal of Infectious Diseases. 2020;101:403–4. doi: 10.1016/j.ijid.2020.09.1059.

13. Cho S, Jackson CR, Frye JG. Freshwater environment as a reservoir of extended-spectrum β-lactamase-producing Enterobacteriaceae. Journal of Applied Microbiology. 2023;134(3):lxad034. doi: 10.1093/jambio/lxad034.

14. Rigby J, Elmerhebi E, Diness Y, Mkwanda C, Tonthola K, Galloway H, et al. Optimised methods for detecting Salmonella Typhi in the environment using validated field sampling, culture, and confirmatory molecular approaches. Journal of applied microbiology. 2021. Epub 2021/07/30. doi: 10.1111/jam.15237. PubMed PMID: 34324765.

15. Tanner WD, Atkinson RM, Goel RK, Toleman MA, Benson LS, Porucznik CA, et al. Horizontal transfer of the blaNDM-1 gene to Pseudomonas aeruginosa and Acinetobacter baumannii in biofilms. FEMS microbiology letters. 2017;364(8). Epub 2017/03/24. doi: 10.1093/femsle/fnx048. PubMed PMID: 283332341.

16. Beck NK, Callahan K, Nappier SP, Kim H, Sobsey MD, Meschke JS. Development of a spot-titer culture assay for quantifying bacteria and viral indicators. Journal of Rapid Methods & Automation in Microbiology. 2009;17(4):455–64. doi: 10.1111/j.1745-4581.2009.00182.x.

17. Mazumder R, Abdullah A, Ahmed D, Hussain A. High Prevalence of bla(CTX-M-15) Gene among Extended-Spectrum β-Lactamase-Producing Escherichia coli Isolates Causing Extraintestinal Infections in Bangladesh. Antibiotics (Basel, Switzerland). 2020;9(11). Epub 2020/11/15. doi: 10.3390/antibiotics9110796. PubMed PMID: 33187055; PubMed Central PMCID: PMCPMC7696227.

18. Woodford N, Fagan EJ, Ellington MJ. Multiplex PCR for rapid detection of genes encoding CTX-M extended-spectrum (beta)-lactamases. The Journal of antimicrobial chemotherapy. 2006;57(1):154–5. Epub 2005/11/15. doi: 10.1093/jac/dki412. PubMed PMID: 16284100.

19. National Institute of Health (NIH) Islamabad. Weekly Field Epidemiological Report July 4-10, 2021. Field Epidemiology & Disease Surveillance Division; 2021 July 14, 2021. Report No.

20. National Institute of Health (NIH) Islamabad. Weekly Field Epidemiological Report August 15-21, 2021. Field Epidemiology & Disease Surveillance Division; 2021 November 11, 2020. Report No.

21. Dos Santos RL, Davanzo EFA, Palma JM, Castro VHL, da Costa HMB, Dallago BSL, et al. Molecular characterization and biofilm-formation analysis of Listeria monocytogenes, Salmonella spp., and Escherichia coli isolated from Brazilian swine slaughterhouses. PloS one. 2022;17(9):e0274636. Epub 2022/09/21. doi: 10.1371/journal.pone.0274636. PubMed PMID: 36126071; PubMed Central PMCID: PMCPMC9488830.

22. U.S. Environmental Protection Agency. Health Risks from Microbial Growth and Biofilms in Drinking Water Distribution Systems In: Water OoGWaD, editor. Washington DC 2002.

23. Ghafourian S, Sadeghifard N, Soheili S, Sekawi Z. Extended Spectrum Beta-lactamases: Definition, Classification and Epidemiology. Current issues in molecular biology. 2015;17:11–21. Epub 2014/05/14. PubMed PMID: 24821872.

24. Ahsan A, Rehman TAU, Irshad H, Shahzad MA, Siddique A, Jamil A, et al. Antibiotic resistance pattern and molecular detection of ESBL-associated genes in E. coli from surface and wastewater of Islamabad capital territory, Pakistan. Journal of water and health. 2022;20(4):601–9. Epub 2022/04/29. doi: 10.2166/wh.2022.216. PubMed PMID: 35482377.

25. Nga TV, Karkey A, Dongol S, Thuy HN, Dunstan S, Holt K, et al. The sensitivity of real-time PCR amplification targeting invasive Salmonella serovars in biological specimens. BMC infectious diseases. 2010;10:125. Epub 2010/05/25. doi: 10.1186/1471-2334-10-125. PubMed PMID: 20492644; PubMed Central PMCID: PMCPMC2886058.

